# Continuous Glucose Monitoring Improves Detection of Clinically Significant Dysglycemia in Hospitalized Patients With Type 2 Diabetes or Hyperglycemia: A Prospective Real-World Study

**DOI:** 10.64898/2026.06.27.26356759

**Authors:** Hector Del Rio Zanatta, Luis Montiel Lopez, Leticia Lopez Carreola, Alexis Zambrano Zambrano, Kevin Zambrano Zambrano, Brian Bernal Alferes, Fernando Diaz Basilio, Angel Alfonso Garduño Pérez

**Affiliations:** Centro Medico Nacional 20 de Noviembre, ISSSTE, Mexico City, Mexico

**Keywords:** continuous glucose monitoring, inpatient diabetes, dysglycemia, hypoglycemia, hyperglycemia, time in range, hospital glycemic control

## Abstract

**Background:** Continuous glucose monitoring (CGM) is increasingly used for inpatient glycemic surveillance, but evidence in non-critical care wards remains limited, particularly in real-world public healthcare settings. Intermittent capillary glucose testing may fail to detect transient, nocturnal, or asymptomatic dysglycemia.

**Objective:** To evaluate whether CGM improves detection of clinically significant dysglycemia compared with seven-point capillary glucose monitoring in hospitalized patients with type 2 diabetes mellitus or hyperglycemia.

**Methods:** Prospective, observational, non-randomized, real-world study performed in a tertiary referral center in Mexico. Fifty-six hospitalized patients were included: 28 underwent flash CGM and 28 underwent seven-point capillary glucose monitoring. Patients were followed for up to 6 hospitalization days. The main analytical focus was detection of clinically significant dysglycemia, including hypoglycemia <70 mg/dL, clinically significant hypoglycemia <54 mg/dL, and severe hyperglycemia >250 mg/dL. Secondary outcomes included time in range, mean daily glucose, insulin requirements, infectious complications, length of stay, and mortality.

**Results:** CGM detected more hypoglycemia <70 mg/dL than capillary monitoring (71.4% vs 35.7%, p=0.005), more clinically significant hypoglycemia <54 mg/dL (median 3 [IQR 0-6.5] vs 0, p=0.030), and more severe hyperglycemia >250 mg/dL (median 8.5 [IQR 0.5-17] vs 0 [IQR 0-9.52], p=0.030). Time in range was not significantly different between groups (59.86 +/- 23.46% vs 69.28 +/- 24.99%, p=0.151). After adjustment for age, diabetes duration, and admission hyperglycemia, CGM remained associated with hypoglycemia detection (OR 4.7, 95% CI 1.2-19.0, p=0.027).

**Conclusions:** In this prospective real-world inpatient study, CGM improved detection of clinically significant dysglycemia during up to 6 hospitalization days. Although CGM did not improve time in range or short-term clinical outcomes, it provided superior glycemic surveillance compared with intermittent capillary glucose testing.

## Introduction

Glycemic control is a core component of inpatient care for patients with type 2 diabetes mellitus and for patients who develop hyperglycemia during hospitalization. Both hyperglycemia and hypoglycemia have been associated with adverse outcomes, including infectious complications, prolonged hospitalization, cardiovascular events, and mortality. (1,2)

Conventional capillary glucose monitoring remains the most widely used method for inpatient glycemic assessment. However, intermittent testing provides isolated glucose values at predetermined times and may fail to identify transient postprandial hyperglycemia, nocturnal hypoglycemia, asymptomatic hypoglycemia, and clinically relevant glycemic variability.

Continuous glucose monitoring provides a more complete assessment of glucose behavior by capturing trends and glycemic excursions throughout the day. In addition to glucose values, CGM allows evaluation of time in range, time below range, time above range, and variability metrics. These parameters are increasingly recognized as clinically relevant complements to traditional glycemic indicators. (4)

Despite strong adoption in outpatient diabetes care, the role of CGM in hospitalized non-critical patients remains under investigation. Available studies suggest that CGM may detect substantially more hypoglycemic events than point-of-care capillary testing, but whether this translates into improved clinical outcomes remains uncertain. (4-6)

The present study evaluates CGM implementation in a real-world tertiary-care public hospital setting in Mexico. Rather than positioning CGM solely as a strategy to improve mean glucose or time in range, this study focuses on its value as a tool for improved inpatient glycemic surveillance and detection of clinically significant dysglycemia.

## Methods

### Study design and setting

This was a prospective, observational, non-randomized real-world study conducted at Centro Medico Nacional 20 de Noviembre, ISSSTE, a tertiary referral center in Mexico City. The study was performed between June and November 2024.

### Study population

Hospitalized adult patients with type 2 diabetes mellitus or hyperglycemia during hospitalization were consecutively included. Patients were monitored according to institutional clinical practice and device availability during the study period.

### Monitoring strategies

The CGM group underwent flash continuous glucose monitoring. The comparison group underwent intermittent seven-point capillary glucose monitoring using reagent strips, performed fasting, 2 hours after breakfast, before lunch, 2 hours after lunch, before dinner, 2 hours after dinner, and at 03:00 am.

### Follow-up

Patients were followed for a maximum of 6 hospitalization days. Glycemic data and clinical outcomes were collected during this monitoring period.

### Outcomes

The main analytical focus was detection of clinically significant dysglycemia, including hypoglycemia <70 mg/dL, clinically significant hypoglycemia <54 mg/dL, and severe hyperglycemia >250 mg/dL. Secondary outcomes included time in range (70-180 mg/dL), mean daily glucose, insulin requirements, infectious complications, overall complications, length of hospital stay, and in-hospital mortality.

### Statistical analysis

Continuous variables were assessed for distribution and summarized as mean +/- standard deviation or median and interquartile range. Categorical variables were summarized as frequencies and percentages. Comparisons were performed using Student’s t-test, Mann-Whitney U test, chi-square test, or Fisher’s exact test, as appropriate. Binary logistic regression models were used to estimate odds ratios with 95% confidence intervals. Adjusted models included baseline variables that differed between groups, including age, diabetes duration, and admission due to hyperglycemic decompensation. A two-sided p value <0.05 was considered statistically significant.

### Ethical considerations

The study was conducted according to the Declaration of Helsinki and approved by the institutional ethics committee. All participants provided informed consent according to institutional requirements.

## Results

A total of 56 patients were included: 28 in the CGM group and 28 in the seven-point capillary glucose monitoring group. Patients were followed during up to 6 hospitalization days.

Baseline characteristics are shown in Table 1. Both groups had identical sex distribution, with 18 men (64.3%) and 10 women (35.7%) in each group. Patients in the CGM group were younger than those in the capillary monitoring group (56 +/- 11 vs 65 +/- 11.72 years, p=0.004). Diabetes duration was also shorter in the CGM group (median 10 [IQR 5.25-18.75] vs 20 [IQR 12-22] years, p=0.019). Admission due to uncontrolled hyperglycemia was more frequent in the CGM group (50% vs 21.4%, p=0.026).

**Table 1.** Baseline characteristics.

| Variable | CGM n=28 | CG n=28 | p value |
| --- | --- | --- | --- |
| Male | 18 (64.3%) | 18 (64.3%) | 0.613 |
| Female | 10 (35.7%) | 10 (35.7%) |  |
| Age, years | 56 ± 11 | 65 ± 11.72 | 0.004 |
| Time to diabetes diagnosis, years | 10 (5.25-18.75) | 20 (12-22) | 0.019 |
| Reason for admission |  |  |  |
| Uncontrolled hyperglycemia | 14 (50%) | 6 (21.4%) | 0.026 |
| Infectious process | 7 (25%) | 13 (46.4%) | 0.094 |
| Cardiac disease / CHF | 2 (7.1%) | 0 | 0.245 |
| Oncologic disease | 4 (14.2%) | 1 (3.57%) | 0.176 |
| Upper gastrointestinal bleeding | 0 | 3 (10.7%) | 0.118 |
| Other causes | 1 (3.5%) | 5 (17.8%) | 0.096 |
| Previous treatment |  |  |  |
| Use of OADs | 15 (53.5%) | 20 (71.4%) | 0.287 |
| DPP-4 inhibitors | 7 (25%) | 10 (35.7%) | 0.383 |
| Metformin | 6 (21.4%) | 7 (25%) | 0.753 |
| SGLT2 inhibitors | 3 (10.7%) | 2 (7.1%) | 0.530 |
| HbA1c at admission, % | 9.61 +/- 2.26 | 8.72 +/- 2.34 | 0.191 |

Values are expressed as frequency (%), mean +/- SD, or median (IQR). CGM: continuous glucose monitoring; CG: capillary glucose; CHF: congestive heart failure; OADs: oral antidiabetic drugs.

Clinical outcomes and glycemic metrics are summarized in Table 2. CGM identified more patients with hypoglycemia <70 mg/dL than capillary glucose monitoring (71.4% vs 35.7%, p=0.005). The median number of clinically significant hypoglycemia events <54 mg/dL was also higher with CGM (3 [IQR 0-6.5] vs 0, p=0.030). Severe hyperglycemia >250 mg/dL was more frequently detected with CGM (8.5 [IQR 0.5-17] vs 0 [IQR 0-9.52], p=0.030).

**Table 2.** Clinical outcomes and glycemic control.

| Variable | CGM n=28 | CG n=28 | p value |
| --- | --- | --- | --- |
| Complications | 14 (50%) | 17 (60.7%) | 0.296 |
| Infectious process | 12 (42.9%) | 19 (67.8%) | 0.060 |
| Hypoglycemia <70 mg/dL | 20 (71.4%) | 10 (35.7%) | 0.005 |
| Length of stay, days | 6 (4-10.5) | 5.5 (3-10) | 0.382 |
| Mortality | 0 | 1 (1.8%) | 0.530 |
| Mean daily glucose - Day 1, mg/dL | 168.6 +/- 66.46 | 160.2 +/- 48.53 | 0.587 |
| Mean daily glucose - Day 2, mg/dL | 156.7 +/- 64.36 | 150.6 +/- 47.38 | 0.692 |
| Mean daily glucose - Day 3, mg/dL | 152.2 +/- 44.95 | 158.0 +/- 61.01 | 0.702 |
| Mean daily glucose - Day 4, mg/dL | 152.8 +/- 49.08 | 148.5 +/- 50.31 | 0.807 |
| Mean daily glucose - Day 5, mg/dL | 139.6 +/- 45.15 | 162.6 +/- 39.17 | 0.211 |
| Mean daily glucose - Day 6, mg/dL | 138.5 +/- 40.48 | 160.2 +/- 43.94 | 0.308 |
| Low glucose, 54-69 mg/dL | 2.5 (0-8) | 0 (0-4.76) | 0.101 |
| Very low glucose, <54 mg/dL | 3 (0-6.5) | 0 | 0.030 |
| Hyperglycemia >180 mg/dL | 26 (92.9%) | 22 (78.6%) | 0.384 |
| High glucose, 181-250 mg/dL | 21 (8-28) | 14.2 (3.81-35.7) | 0.755 |
| Very high glucose, >250 mg/dL | 8.5 (0.5-17) | 0 (0-9.52) | 0.030 |
| Time in range, 70-180 mg/dL, % | 59.86 +/- 23.46 | 69.28 +/- 24.99 | 0.151 |
| Total insulin, units | 151 (86.5-247) | 83 (31-154.5) | 0.004 |

Values are expressed as frequency (%), mean +/- SD, or median (IQR). Glycemic events were evaluated during up to 6 hospitalization days. CGM: continuous glucose monitoring; CG: capillary glucose.

Mean daily glucose values across 6 hospitalization days were not significantly different between groups. Time in range was numerically lower in the CGM group but not statistically different (59.86 +/- 23.46% vs 69.28 +/- 24.99%, p=0.151). Total insulin use was higher in the CGM group (median 151 [IQR 86.5-247] vs 83 [IQR 31- 154.5] units, p=0.004).

Regression models are shown in Table 3. CGM was associated with hypoglycemia detection in the unadjusted model (OR 3.6, 95% CI 1.2-11.2, p=0.024) and remained associated after adjustment for age, diabetes duration, and admission hyperglycemia (OR 4.7, 95% CI 1.2-19.0, p=0.027).

**Table 3.** Logistic regression models.

| Outcome | Unadjusted OR (95% CI) | Adjusted OR (95% CI) | p value adjusted |
| --- | --- | --- | --- |
| Hypoglycemia detection | 3.6 (1.2-11.2), p=0.024 | 4.7 (1.2-19.0) | 0.027 |
| Infectious process | 0.3 (0.10-0.91) p=0.035 | 0.3 (0.06-1.4) | 0.125 |
| Prolonged hospital stay | 1.38 (0.45-4.28) p=0.568 | 4.98 (0.7-24.1) | 0.121 |

Adjusted model included age, diabetes duration, and admission due to hyperglycemic decompensation. OR: odds ratio; CI: confidence interval.

## Figures

**Figure 1.**
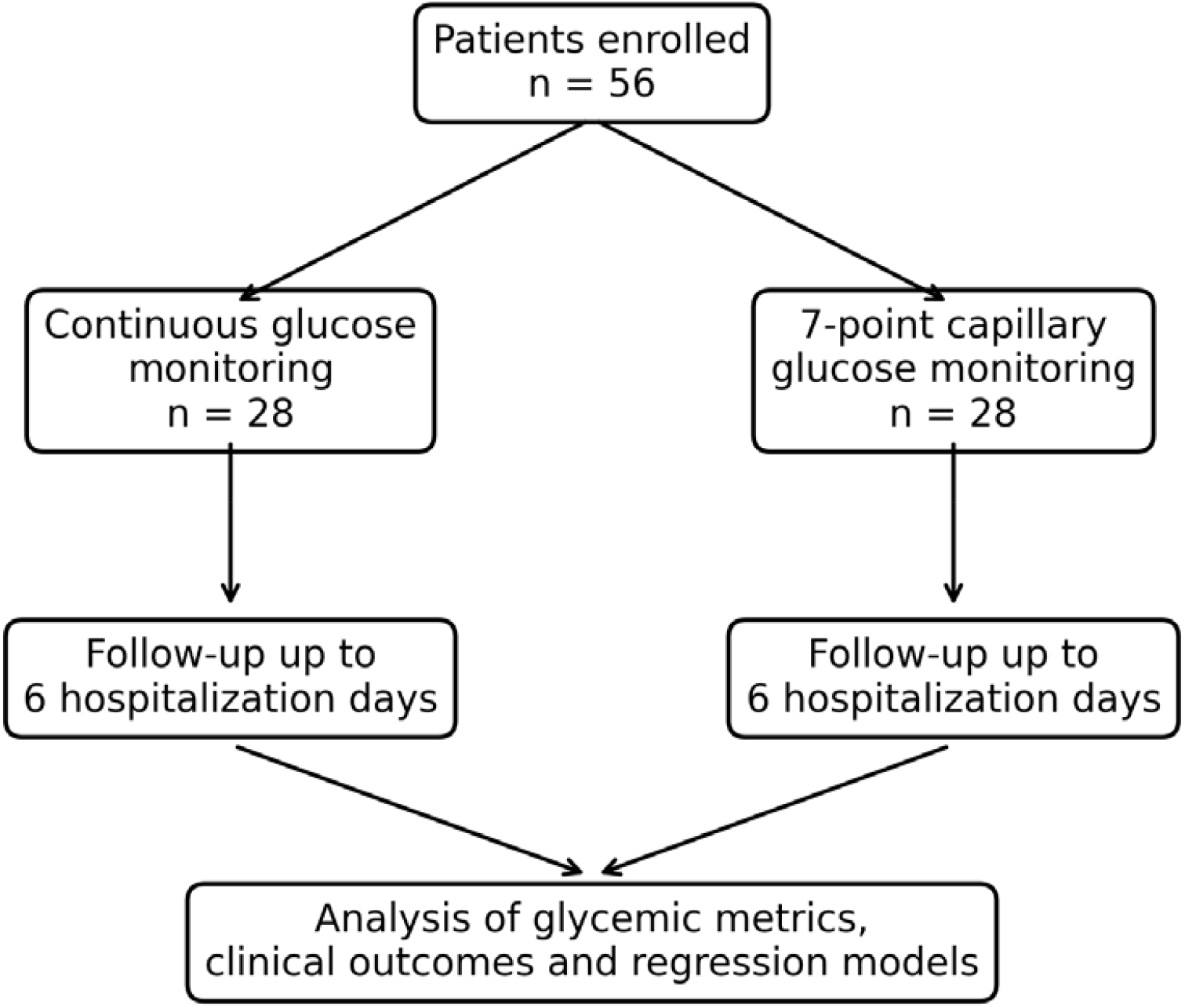
Study flowchart.

**Figure 2.**
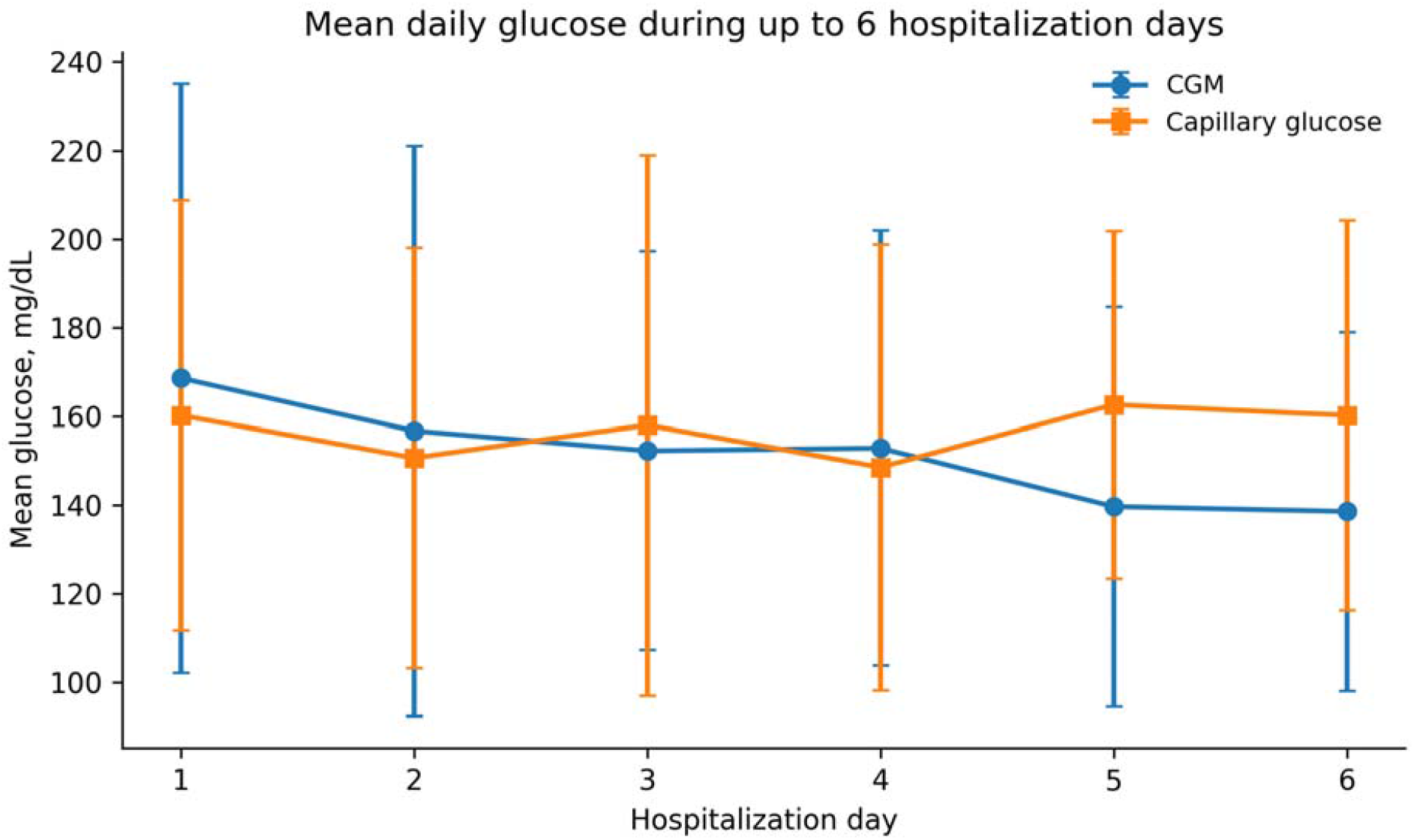
Mean daily glucose during up to 6 hospitalization days. Error bars represent SD.

**Figure 3.**
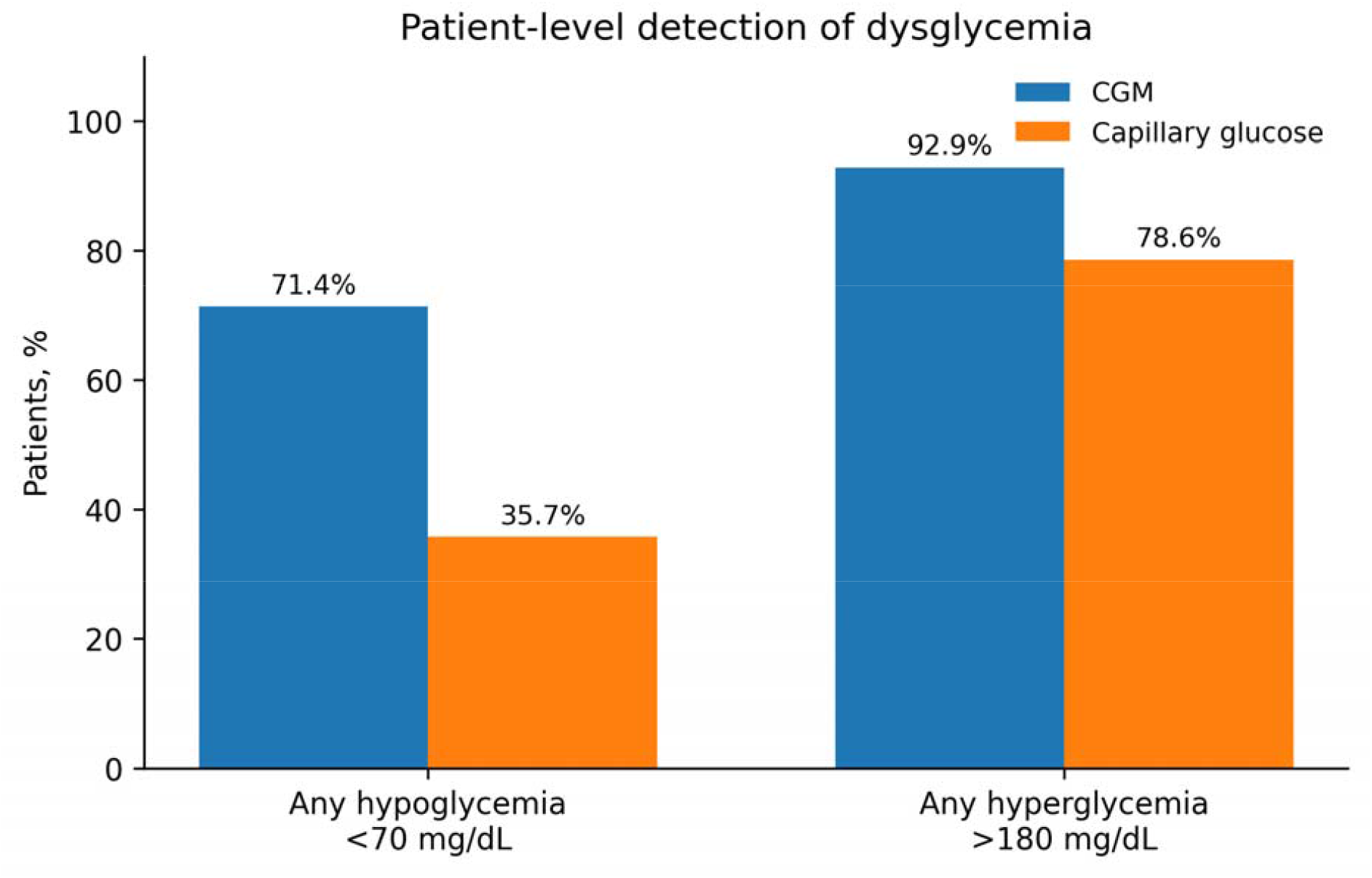
Patient-level detection of dysglycemia according to monitoring strategy.

**Figure 4.**
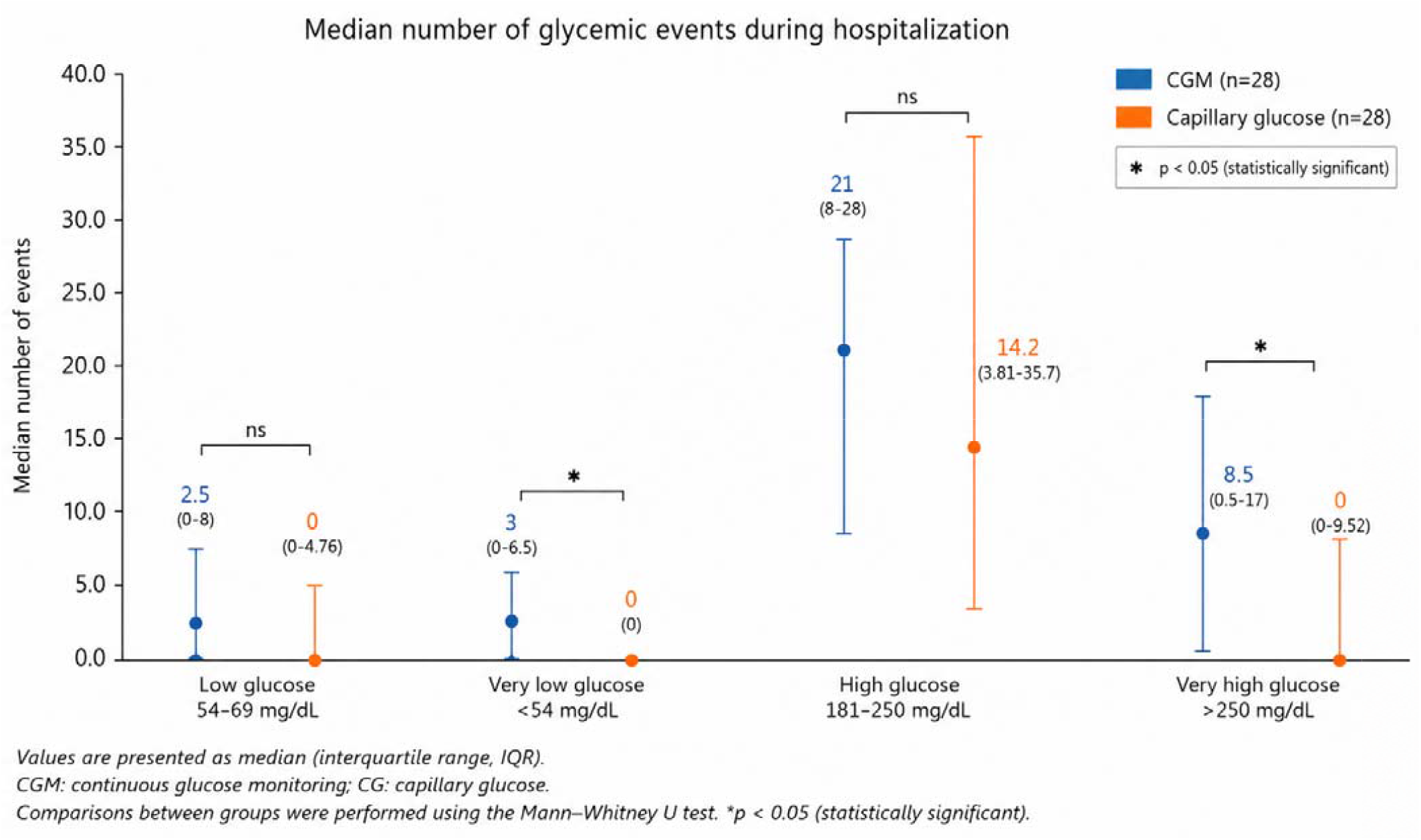
Median number of glycemic events during hospitalization.

**Figure 5.**
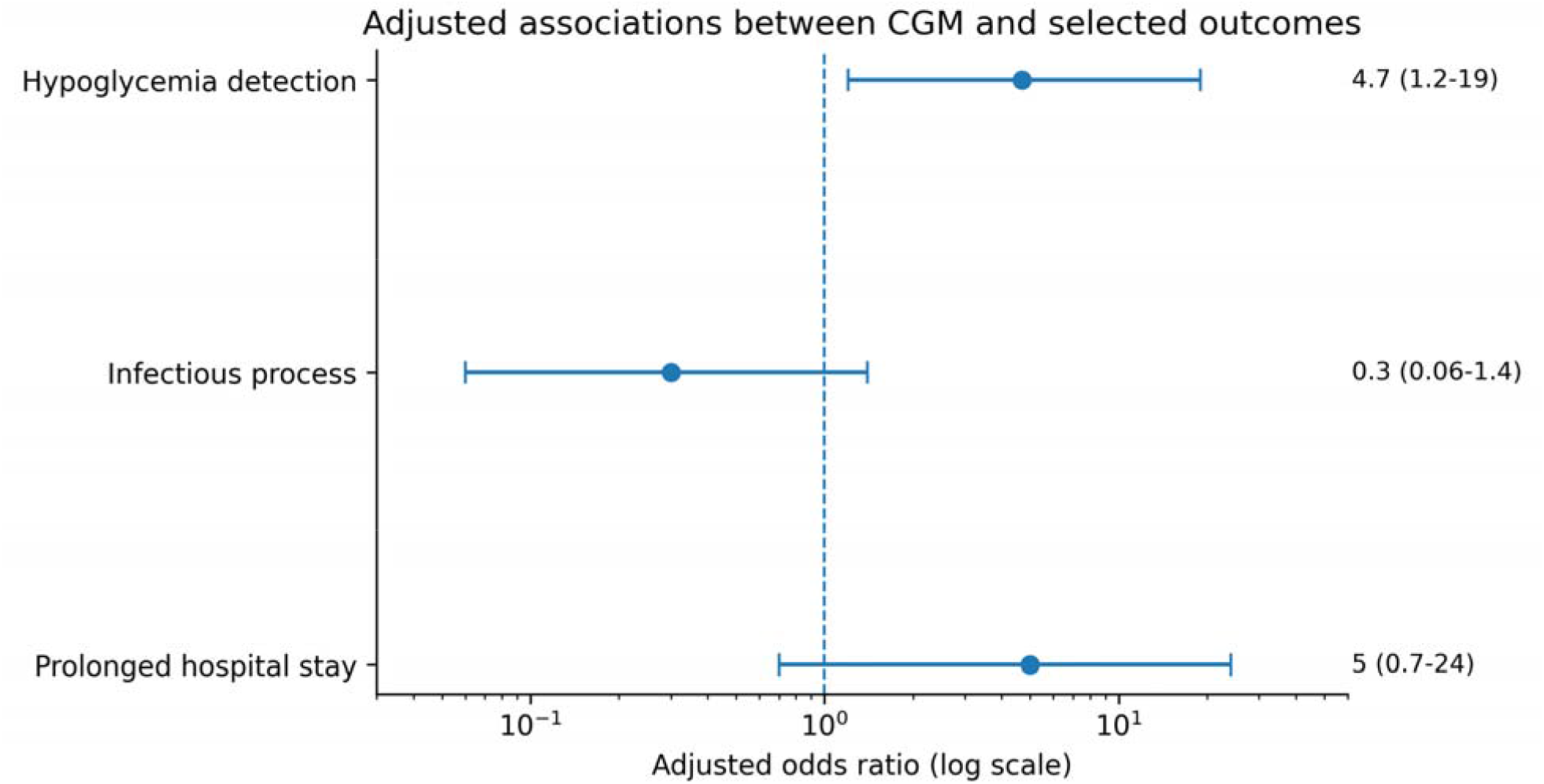
Adjusted associations between CGM and selected outcomes.

## Discussion

This prospective real-world study found that CGM improved detection of clinically significant dysglycemia in hospitalized patients with type 2 diabetes mellitus or hyperglycemia. The most relevant finding was not an improvement in time in range, but rather improved identification of hypoglycemia, clinically significant hypoglycemia, and severe hyperglycemia during up to 6 hospitalization days.

This distinction is important. Intermittent capillary monitoring provides a limited number of measurements and may underestimate glycemic variability. Therefore, the higher frequency of events detected in the CGM group should be interpreted primarily as improved detection sensitivity rather than a true increase in dysglycemia caused by CGM.

The increased detection of hypoglycemia is clinically relevant because inpatient hypoglycemia is associated with arrhythmias, neurological injury, falls, cardiovascular complications, and mortality. Severe hypoglycemia may be especially difficult to detect when it occurs overnight or without symptoms. CGM may therefore improve patient safety by revealing events that conventional monitoring misses. (1,4,6,7)

A similar interpretation applies to severe hyperglycemia. Although the proportion of patients with hyperglycemia>180 mg/dL was similar between groups, CGM detected more severe hyperglycemia >250 mg/dL. This finding suggests that CGM better characterizes glycemic variability and extreme excursions, which may support more precise therapeutic decisions.

Time in range did not significantly differ between groups. This negative primary glycemic control finding should not be interpreted as evidence that CGM lacks value in the hospital. Rather, it indicates that, in this study, CGM functioned mainly as a superior surveillance tool instead of producing measurable short-term improvement in average glycemic control.

Total insulin requirements were higher in the CGM group. This may reflect more frequent recognition of hyperglycemic excursions and subsequent treatment adjustments. However, this finding should be interpreted cautiously due to the observational design, baseline imbalance, and potential confounding by indication.

No significant differences were observed in mortality, infectious complications, or length of stay. The study was not powered to detect differences in hard clinical outcomes, and the sample size limits conclusions regarding these endpoints. Larger multicenter randomized studies are required to determine whether improved dysglycemia detection translates into better clinical outcomes. (5,6)

The study has several strengths, including prospective data collection, direct comparison of monitoring strategies, and implementation in a real-world tertiary-care public hospital. This context is relevant because inpatient CGM data from Latin American public healthcare systems remain limited.

Limitations include non-randomized allocation, small sample size, baseline differences between groups, and lack of systematic capillary confirmation for every CGM-detected event. Residual confounding cannot be excluded despite multivariable adjustment.

Overall, the findings support reframing inpatient CGM as a tool for glycemic surveillance and safety rather than solely as an intervention to improve time in range.

## Conclusion

Continuous glucose monitoring improved detection of clinically significant hypoglycemia and severe hyperglycemia during hospitalization, supporting its role as a superior inpatient glycemic surveillance strategy compared with intermittent seven-point capillary glucose testing. Although no significant differences were observed in time in range or short-term clinical outcomes, CGM may facilitate earlier recognition of glycemic excursions and safer inpatient diabetes management.

## Data Availability

All data produced in the present work are contained in the manuscript

## References

1. Korytkowski MT, Muniyappa R, Antinori-Lent K, Donihi AC, Drincic AT, Hirsch IB, et al. Management of hyperglycemia in hospitalized adult patients in noncritical care settings: an Endocrine Society clinical practice guideline. J Clin Endocrinol Metab. 2022;107(8):2101–2128. doi:10.1210/clinem/dgac278.

2. Joseph JJ, Deedwania P, Acharya T, Aguilar D, Bhatt DL, Chyun DA, et al. Comprehensive management of cardiovascular risk factors for adults with type 2 diabetes: a scientific statement from the American Heart Association. Circulation. 2022;145(9):e722–e759. doi:10.1161/CIR.0000000000001040.

3. Wu Z, Liu J, Zhang D, Kang K, Zuo X, Xu Q, et al. Expert consensus on the glycemic management of critically ill patients. J Intensive Med. 2022;2(3):131–145. doi:10.1016/j.jointm.2022.05.004.

4. Yoo HJ, An HG, Park SY, Ryu OH, Kim HY, Seo JA, et al. Continuous glucose monitoring in the hospital. Endocrinol Metab (Seoul). 2021;36(2):240–255. doi:10.3803/EnM.2021.1049.

5. Gómez AM, Umpierrez GE, Muñoz OM, et al. Continuous glucose monitoring versus capillary point-of-care testing for inpatient glycemic control in type 2 diabetes patients hospitalized in the general ward and treated with a basal-bolus insulin regimen. J Diabetes Sci Technol. 2015;10(2):325–329. doi:10.1177/1932296815592425.

6. Galindo RJ, Migdal AL, Davis GM, et al. Comparison of the FreeStyle Libre Pro flash continuous glucose monitoring system and point-of-care capillary glucose testing in hospitalized patients with type 2 diabetes treated with a basal-bolus insulin regimen. Diabetes Care. 2020;43(11):2730–2735. doi:10.2337/dc20-0845.

7. Spanakis EK, Levitt DL, Siddiqui T, et al. The effect of continuous glucose monitoring in preventing inpatient hypoglycemia in general wards. Diabetes Technol Ther. 2018;20(11):753–760.

